# Associations Between Early-Life Indoor and Outdoor Air Pollution Exposure and Childhood Asthma

**DOI:** 10.64898/2026.09.23.26363754

**Authors:** Michael Forsmann, Mario Lovrić, Frederikke Rosenvinge Skov, Olalekan Akintande, Jakob Stokholm, Klaus Bønnelykke, Ann-Marie Malby Schoos, Morten Arendt Rasmussen

## Abstract

**Background:** Early-life air pollution exposure has been associated with childhood asthma, but its relative contribution is poorly characterised in cohorts with repeated indoor measurements. Short sampling windows and seasonal bias limit how well any single measurement can represent cumulative exposure.

**Objective:** To evaluate the association between early-life indoor and outdoor pollutant exposure from birth to 18 months and asthma development at ages 7, 12, and 18 years, while testing potential exposure-exposure and gene-environment interactions.

**Methods:** In the COPSAC**_2000_** birth cohort (***n* = 411**), indoor PM**_2.5_**, black carbon, NO**_2_**, NO***_x_*** and VOCs, and modelled outdoor PM**_2.5_**, NO**_2_** and NO***_x_***, were assessed at 6, 12 and 18 months of age. Indoor metrics were corrected for pump-flow drift and seasonal bias via a Gaussian process with a periodic kernel. Hierarchical Bayesian logistic regression(HBLR) and calibrated random forests (CRF) were evaluated using stratified nested cross-validation for discrimination using AUC-ROC, Calibration using integrated calibration index (ICI), and Brier skill scores. Exposure-exposure and gene-environment interactions were quantified via Shapley interaction indices.

**Results:** Discrimination was weak at all endpoints with test AUC-ROC: 0.58-0.67, BSS near zero (0.012 to 0.05) and ICI: 0.043-0.061 for random forest and AUC-ROC 0.53-0.61, BSS equal to zero and ICI: 0.054-0.067 for hierarchical Bayesian logistic regression (HBLR). No pollutant reached a highest density interval excluding OR **= 1**. The polygenic risk score(PRS) was the only term non-null at all three ages in the adjusted models. While gene-environment interactions were detected, effect sizes were negligible and statistically indistinguishable from zero.

**Significance:** Where ambient pollution is low, cumulative early-life exposure adds little to long-term asthma prediction beyond genetic and contextual factors, and an improved exposure correction does not uncover an effect.

**Impact:** Repeated bedroom measurements across infancy are rare, and short sampling windows make them hard to read as cumulative exposure. We corrected measured concentrations for pump-flow drift and for seasonal sampling bias on the pollutants and used two complementary machine learning models to predict interaction and risk. In this low-exposure Danish cohort, genetic risk and residential context dominated, and the measured pollutants added almost nothing to the asthma risk at 7,12 and 18 years. These null results indicate that early cumulative exposures of indoor and outdoor air pollution contribute little to nothing to asthma up to 18 years.

## Introduction

Asthma is the most prevalent chronic respiratory disease worldwide. The Global Burden of Disease Study estimated 262 million prevalent cases in 2019 (95% UI 224-309 million) [1]. Estimates are, however, highly sensitive to the case definition applied: global prevalence in those aged 5-69 years ranges from 5.4% for current asthma to 11.5% for current wheezing, reflecting the absence of a universally accepted definition or confirmatory test [2]. Prevalence and incidence are higher in high socio-demographic index (SDI) settings, whereas mortality and disability-adjusted life years are concentrated in lower SDI quintiles [1], reflecting differences in diagnosis, case ascertainment and access to treatment as well as in underlying risk. This heterogeneity in ascertainment motivates the use of prospectively collected, physician-confirmed diagnoses when studying aetiology. The cause is likely multifactorial, arising from a complex interplay between genetics, immune stimulation, and environmental exposures that are particularly influential during the first years of life [3]. Children may be particularly susceptible to airborne pollutants because of their developing physiology, including immature lungs and a developing immune system [4, 5]. Furthermore, higher ventilation rates relative to body weight, along with underdeveloped detoxification pathways, can potentially intensify adverse effects of exposure to suboptimal air quality [6, 7]. Children spend most of their time indoors - not only at home but also in kindergartens, schools, and daycare facilities [8], making indoor air quality (IAQ) a key determinant of cumulative pollutant exposure. Early-life exposure to airborne pollutants is widely recognised as a major risk factor for asthma and related clinical traits including wheeze and lung function, with numerous epidemiological studies showing associations with fine particulate matter (PM_2.5_), black carbon, nitrogen oxides (NO*_x_*, NO_2_) and volatile organic compounds ( VOCs) [9–12]. However, the strength of these associations varies across cohorts, partly due to limitations in exposure assessment, which often relies on short-term measurements or inferred outdoor concentrations, and on inevitable confounding by social determinants and geographic location. Further, methods for quantifying IAQ frequently fail to capture temporal variability and lack personal exposure data, both of which are essential for understanding individual risk [13]. As a result, the biological mechanisms underlying these associations remain incompletely characterised. Recent studies indicate that genetics predicts asthma independently of conventional risk factors [14]. However, the extent to which genetic risk is modulated by environmental exposures remains uncertain [15]. Gene-environment (G · E) interactions for pollutants are plausible, as pollutants can intensify airway inflammation [16–19], oxidative stress and immune dysregulation, particularly in genetically susceptible individuals. Demonstrating such interactions would help explain why some children develop asthma despite low exposure, while others remain unaffected despite high pollutant levels [16, 18–20]. The relative contributions of outdoor and indoor pollution sources, as well as their interactions with covariates, are rarely characterised within a longitudinal framework. The Copenhagen Prospective Studies on Asthma in Childhood (COPSAC_2000_) birth cohort provides a unique opportunity to address these research gaps. With early-life longitudinal measures of indoor exposures (PM_2.5_, black carbon, NO*_x_*, NO_2_, formaldehyde, acetaldehyde, and acetone) at 6, 12, and 18 months of age [21, 22], modelled outdoor concentrations from the same period for NO_2_, NO*_x_*, and PM_2.5_, and clinical prospective follow-up for asthma diagnosis up to 18 years of age, we aim to investigate the association between early life indoor and outdoor air quality and asthma development.

We hypothesised that

1. Cumulative indoor and outdoor pollutant exposures during the first 18 months increase the risk of asthma later in life.
2. There is an interaction between asthma genetics and pollutants, and between exposures, which increases the risk of childhood asthma.

To test these hypotheses, we developed a correction method based on Gaussian processes [23, 24] to mitigate seasonal variation and sampling biases, so that the measurement reflects cumulative exposure rather than the sampling period alone. After the corrections a dual modelling strategy was used: a hierarchical Bayesian logistic regression(HBLR), which provides interpretable posterior odds ratios and credible intervals, and a random forest classifier using Sigmoid calibration, which captures non-linear, higher-order interactions [25].

## Materials and Methods

### Study Population

This study uses data from the COPSAC_2000_ cohort, a prospective, single-centre clinical birth cohort comprising children born between 1998 and 2001 to mothers with diagnosed asthma [26]. The children were seen by COPSAC physicians every 6 months from age 0 to 7 years and at follow-up visits at ages 12 and 18 years. Study design is described in detail elsewhere [26].

### Polygenic risk score

Genetic susceptibility was quantified using a polygenic score for childhood-onset asthma [27]. Asthma has partially distinct genetic architectures depending on age of onset: in a genome-wide analysis of 37,846 asthma cases and 318,237 healthy controls, Pividori et al. [27] identified 61 independent asthma-associated variants and partitioned these into 23 loci specific to childhood-onset disease, 37 loci shared between childhood- and adult-onset disease, and one locus specific to adult-onset disease, with childhood onset defined as diagnosis before 12 years of age. Because all endpoints in the present study are of childhood and adolescent onset, we used the 23 childhood-onset-specific variants to construct a polygenic risk score.

### Asthma Diagnosis and LRTI

Asthma was diagnosed and managed exclusively by COPSAC physicians using a predefined clinical algorithm [26]. The detailed description of the asthma diagnosis can be found in Supplementary S11. Respiratory and gastrointestinal illness episodes were recorded prospectively and reviewed by clinicians at scheduled follow-up visits in COPSAC_2000_ [28].

### Outdoor air pollution

Individual-level maternal and child home addresses from conception through the last day of follow-up, including dates of moving and immigration, were obtained prospectively. The concentrations of ambient PM_2.5_, NO*_x_*, and NO_2_ were modelled at each home address for each week using the Danish Eulerian Hemispheric Model (DEHM)-urban background model, (UBM)-Danish Air Pollution and Human Exposure Modelling System (AirGIS) model system [29–31]. The local contribution from traffic at the street level was calculated by the Operational Street Pollution Model [32]. The local contribution from primary anthropogenic particle sources was calculated at 1 km × 1 km resolution using the UBM [29]. As outdoor exposure, a time-weighted mean of cumulative exposure from birth to 6, 12, and 18 months was used. No more than 10% missing values were allowed for each of the time-weighted means [10, 12].

### Indoor PM_2.5_ and black carbon

PM_2.5_ and black carbon were measured in the children’s bedroom for a duration of 10 days at 6, 12, and 18 months of age. Trained staff installed the sampling device and started and ended the measurements, recording the flow at both events. Measurements were made with a KTL PM_2.5_ cyclone (Korea Testing Laboratory; Jinju-si, South Korea) and a BGI400 pump (BGI Inc., Butler, NJ, USA). Filter reflectance is a measure of the blackness of the PM_2.5_ filters and is used here as an indicator of the black carbon content of the sample [21, 22]. Because the aerodynamic cut-point of the cyclone falls as the pump flow rises, measured concentrations decline systematically with flow; each measurement was therefore rescaled multiplicatively to a nominal flow of 4 L min^−1^ using a Gaussian-process fit of concentration on end-of-sampling flow (Supplementary Eq. (S2)). The correction is monotonic, preserves the ranking of concentrations recorded at a given flow. Full detail is given in Supplementary Sections S1 and S2.

### Indoor NO*_x_*, NO_2_, formaldehyde, acetaldehyde, and acetone

NO*_x_*, NO_2_, formaldehyde, acetaldehyde, and acetone were measured in the bedrooms of the children at 6, 12, and 18 months after birth for approximately 10 weeks. The NO*_x_*, NO_2_, and VOC samplers(Swedish Environmental Research Institute, Gothenburg, Sweden) were provided to the parents, along with detailed instructions for starting and stopping measurements. After 10 weeks, the samplers were returned to the clinical research unit by ordinary mail. The shipping time was short and negligible compared to the 10 week measurement. Details of all measurements can be found in the original papers [21, 22]

Seasonal bias correction is in Supplementary S2.

### Indoor sinks, sources, and home properties

Indoor sink and sources were characterised via parental questionnaire, in which parents reported the number of days per year their child was exposed to each source. Sources included gas stove(source), fireplace(source), carpet(sink), passive smoking(source), and cooker hood operation(sink). These sinks and sources have been previously associated with indoor VOC, particulate, and NO_2_ concentrations in residential settings [9, 33].

Outdoor local sources and sinks were derived from OpenStreetMap extracted within a 5000 m radius of each child’s home address [34]. A 5000 m radius was selected to capture land-use features whose emission plumes or spatial influence extend beyond the immediate residential neighbourhood, including airports, power plants, and coastline, which would be excluded at shorter extraction distances. Sensitivity to the buffer radius was not formally tested. Addresses were recorded at birth and at the first and second years of life; where parents were separated, the exposure estimate was taken as the mean across both residential addresses. Source categories included roads(source), motorways(source), power plants(source), farmland(source), airports(source), and forests(sink), with the extracted area serving as a proxy for each source’s relative emission strength. Home characteristics were retrieved from the Danish Building and Housing Register (BBR) by linking residential addresses to the ‘Datavask’ address-harmonisation function, which returns a validated address identifier and associated building record based on the most likely address. Variables extracted included total area of the house, construction year, and floor level.

### Adjustments and clinical measurements

Primary analyses were performed with adjusted models, including the following pre-specified covariates: BMI at age 6 years (continuous), maternal education at the child’s 1-year visit (4-level ordinal scale: 1 high school, 2 vocational/college, 3 bachelor, 4 master or higher), and two principal components summarising indoor and outdoor pollutant sources and sinks (continuous). PC1 loads mainly on the rural-urban gradient (farmland and forest area against road and built-up area) and PC2 mainly on motorway, coastline and forest area. Correlation to exposures can be seen in Supplementary S3. These covariates were included in all adjusted models. A secondary analysis without the adjustment is in Table 2, and figures of odds ratios, SHAP values and interaction is in Supplementary sections S5 and S6.

### Correction for seasonal sampling bias

Sampling windows were short relative to the year: 10 days for PM_2.5_ and black carbon, and 10 weeks for the VOCs, NO_2_ and NO*_x_*. Each observed concentration therefore largely reflects the season in which it was collected rather than the child’s exposure. Because sampling weeks were distributed across the calendar year, this introduces a nuisance seasonal component into the between-child contrasts.

To account for this, a Gaussian process with a periodic kernel was used to estimate the mean seasonal trend of each pollutant across the year [35]. Writing *G_T_* (·) for the fitted trend of pollutant *T* and *w_i_* for the sampling week of observation *i*, each measurement was rescaled by the ratio of the mean trend over the Δ = 365*/*(7 · 2) weeks preceding the sample to the trend at the sampling week itself:

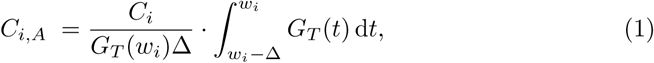

where *C_i_* is the observed concentration, *C_i,A_* the seasonally adjusted concentration, and *t* denotes time/week and *w_i_* the week of the measurement. The integral was evaluated by Simpson’s ⅓ rule on 499 points. The adjusted value therefore estimates the concentration the household would have averaged over the half-year preceding the measurement, rather than during the sampling window alone. Because measurements were scheduled at 6, 12 and 18 months of age, the three half-year windows partition the first 18 months of life without overlap. The unweighted mean of the three adjusted values,

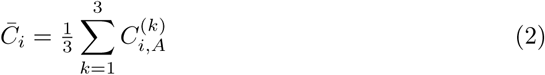

*C_i,A_* adjusted concentration, 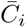 average concentration of the household, k the sample number. The time-weighted average exposure over that period. Where only one measurement was available it was scaled to the whole 18-month period from birth; where two were available, the first was scaled six months forward and backward and the second six months backward, so that no period was counted twice. The correction improved the household-level variance explained for the strongly seasonal pollutants (e.g. NO*_x_*, *R*^2^ from 0.03 to 0.22) and reduced to approximately unity for pollutants with little seasonal structure (Supplementary Section S2).

### Predicting asthma status

Flow- and seasonality-corrected cumulative indoor exposures together with outdoor exposures were used as predictors of asthma at 7, 12, and 18 years. Two complementary models were applied: Hierarchical Bayesian logistic regression (HBLR) and Calibrated random forest(CRF), interpreted with odds ratios and SHAP (Shapley additive explanations) values and interactive values, respectively. The pipeline is summarised in Supplementary Figure S9. Exposures were first corrected for pump-flow drift (Supplementary Eq. (S2)) and then for seasonal sampling bias (Eq. 1). Within each fold, *Z*-normalisation was fitted on the training set and applied to both training and held-out data, followed by uniform-weighted KNN imputation with 25 neighbours (Supplementary Section S4). The data were split twice: an inner loop was used for hyper-parameter selection, and an outer loop provided the performance estimate, ensuring the result does not depend on a single fortunate split. For every chain of the Bayesian model, convergence was checked by requiring *Ȓ <* 1.01 and a truncated effective sample size above 400 for all variables. The full pipeline is shown in Supplementary Section S7.

Performance was assessed with three complementary metrics. The area under the receiver operating characteristic curve (AUC-ROC) quantifies the ability to rank cases above non-cases. The integrated calibration index (ICI) quantifies calibration as the mean absolute difference between predicted probabilities and a Locally Estimated Scatterplot Smoothing(loess)-based estimate of the observed event rate [36, 37]. The Brier Skill Score (BSS) captures both dimensions jointly as the proportional reduction in Brier Score (BS) relative to a prevalence-only baseline [38, 39]:

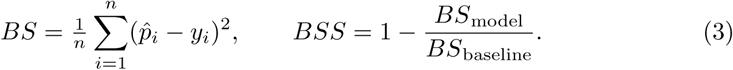

BS is the Brier score, BSS is the Brier skill score. *p^_i_* is the samples predicted chance of having asthma, *y_i_* is the disease outcome 1 sick and 0 healthy of sample i. Reporting all three jointly prevents any one dimension from masking the others [39, 40]. For the HBLR, performance was summarised in two ways. In the pooled evaluation, fold-level predictions used the posterior mean of the coefficients. In the sampled evaluation, predicted probabilities and metrics were recomputed for every posterior draw, propagating parameter uncertainty into the performance estimates and yielding lower but more realistic point estimates with credible intervals.

### Models

The models evaluated are a random forest calibrated with Sigmoid function and a hierarchical Bayesian logistic regression(HBLR). Calibration with Sigmoid regression aligns the random forest’s predicted probabilities with observed event frequencies, so the outputs can be interpreted as risk estimates rather than ranking scores alone [41, 42]. This is an important property when applying tree-based methods to clinical data, since the model should reflect exposure risk rather than only optimise predictive accuracy. The CRF model was trained using 300 estimators. Hyperparameters were optimized via cross-validation within the following intervals: maximum depth (2–6), minimum samples per leaf (6–15), maximum features (0.5–0.7) and maximum samples (0.5–0.7). The HBLR is used for estimating odds ratios and effect sizes, and regularises coefficients toward zero for more realistic estimates

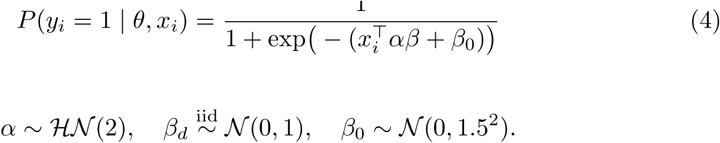

Here *P* (*y_i_* = 1 | *θ, x_i_*) is the probability of a positive outcome for observation *i* given parameters *θ* = (*α, β, β*_0_); *α* is a global shrinkage scale learned from the data; *β* ∈ R*^D^* are the coefficients for the exposure variables; and *β*_0_ is the intercept. The product *αβ* is a non-centred parametrisation of *w_d_* | *α* ∼ *N*(0*, α*^2^), which lets the data determine the overall degree of regularisation.

### Shapley interaction values

Pairwise Shapley interaction indices were computed exactly for tree ensembles with TreeSHAP [43, 44] (tree path dependent). Attributions decompose as *ϕ_i_* = Φ*_ii_* + ∑*_j_*_≠*i*_ Φ*_ij_*, with Φ*_ij_* = Φ*_ji_*, so the pair contributes 2Φ*_ij_*. We report the raw strength 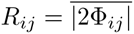 and its size relative to the main effects,

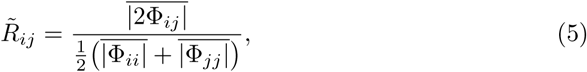

averaged over held-out samples. Both are unsigned, so we add a directional consistency across the *K* folds,

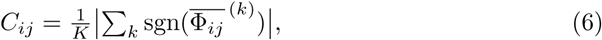

with positive values increasing asthma risk and negative decreasing asthma risk; a real interaction should recur with the same sign in models trained on independent subsets. Pairs were reported only if *C_ij_* ≥ 0.8 and |*ρ_ij_*| ≤ 0.3. The correlation filter is needed because of TreeSHAP conditions on the training distribution along each tree path, so collinear features can be assigned a large apparent Φ*_ij_* even under an additive model.

### Software and LLM

Analysis was performed in Python version 3.14.4. HBLR and inference was implemented using Hamiltonian monte carlo sampler(HMC) and No-U-Turn Sampler(NUTS) in NumPyro 0.21.0[45], and the scaling and the adjusting using pyro using a periodic kernel[46]. The diagnostic parameters and conversion are performed using 1.1.0 Arviz[47]. The correlation statistics are performed with SciPy 1.17.1[48], and the table is made using tableone Python patch version 0.9.6 [49]. BSS, ICI and AUC-ROC were evaluated with scikit-learn 1.8.0 [50], and plots were made with Matplotlib 3.10.9[51] and 1.1.0 Arviz, SHAP plots made with shap 0.51.0[43]. The code and synthetic simulated data can be found on github https://github.com/MichaelForsmann/Predicting-asthma-from-Early-life-Cumulative-indoor-and-outdoor-exposures

A large language model Claude Opus 4.6 was used to assist with revision of text and code. The authors reviewed, verified and take full responsibility for all content.

## Results

Table 1 presents the baseline characteristics of the COPSAC_2000_ cohort stratified by ever-asthma status. The cohort comprised 411 children, of whom 141 (34%) had an asthma diagnosis at least once between 0 and 18 years. BMI at 6 years (*p* = 0.003) and cooker-hood use (*p* = 0.036) were the only variables differing nominally between groups; house area (*p* = 0.083) and PC2 (*p* = 0.060) approached the conventional threshold. These comparisons are unadjusted for multiplicity across 22 tests, and only the BMI difference is significant after Bonferroni correction, so the cooker-hood result should be treated as a chance finding.

**Table 1.**
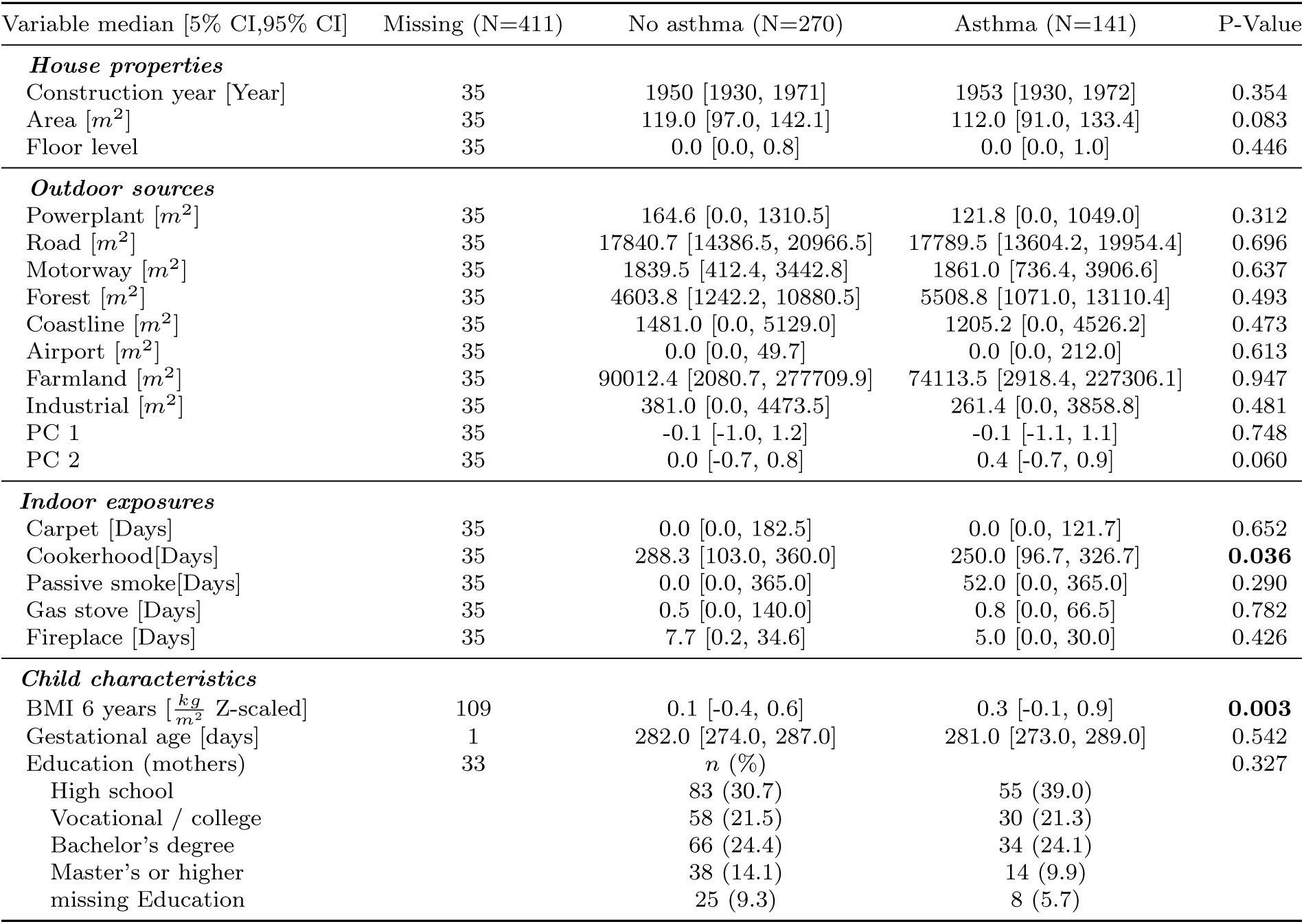
Baseline characteristics of the cohort stratified by ever-asthma status. Continuous variables are presented as median [Q1, Q3] and categorical variables as n (%). P-values are derived using Mann-Whitney U tests for continuous variables and a chi-squared test for education. No correction for multiple comparisons has been applied.

### Distribution of pollutants

Seven pollutants (NO_2_, NO*_x_*, PM_2.5_, black carbon, formaldehyde, acetaldehyde and acetone) were used to quantify indoor air quality, measured as mass per unit volume for all but black carbon, which was quantified by filter reflectance. Formaldehyde was measured at the highest concentrations, with acetaldehyde at approximately one third of those levels. Mean outdoor PM_2.5_ was approximately 62% of the indoor mean, while mean NO_2_ was up to three times higher outdoors than indoors (Supplementary Table S1). Concentrations were affected by season and black carbon and PM_2.5_, pump flow: as pump flow increases the effective cut-point diameter falls and the measured concentration decreases (Supplementary Section S1). The seasonal pattern is broadly sinusoidal, peaking in winter and lowest in summer (Supplementary Figures S2 and S3).

Modelled street-level concentrations in this cohort (median PM_2.5_ 11.9 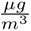, NO2 21.5 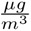; Supplementary Table S1) are low relative to most published birth cohorts but exceed the 2021 WHO annual air quality guideline values of 5 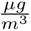 for PM_2.5_ and 10 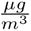 for NO_2_ [52].

### Model performance

To evaluate the performance of models for predicting asthma status at 7, 12 and 18 years of age, AUC-ROC, ICI and BSS are shown in Table 2 for both unadjusted and adjusted models. The Random Forest model achieved test AUC-ROC values ranging from 0.58 to 0.64, outperforming the unadjusted baseline HBLR (test AUC-ROC: 0.53-0.58). The Adjusted Random Forest pipeline reached its peak out-of-fold discrimination at the 7-year endpoint (AUC-ROC = 0.67, 95%CI: [0.54, 0.78]) and at the 12-year endpoint (AUC-ROC = 0.63, 95% CI: [0.54, 0.71]). Correspondingly, the Adjusted HBLR structure achieved minor improvements over its unadjusted counterpart, peaking at an AUC-ROC of 0.61 (95%CI: [0.46, 0.75]) at 12 years. Over-fitting was visible across all tree-based architectures-for instance, the unadjusted Random Forest achieved a training AUC-ROC of 0.90 (95% CI: [0.82, 0.96]) at 12 years but dropped to 0.64 in out-of-fold verification. The out-of-fold test BSS hovered near zero, spanning a narrow interval from -0.010 to 0.050. The Adjusted Random Forest at 7 years exhibited the highest point estimate (BSS = 0.050, 95% CI: [-0.047, 0.11]). Conversely, the HBLR models frequently registered negative mean values on held-out tests ( 18-year unadjusted BSS = -0.010 and 7-year adjusted BSS = -0.008), proving that the models struggled to outperform simple historical probability assignment. The ICI was between 0.043 and 0.067. The adjusted calibrated random forest at 12 years had the lowest calibration error ICI = 0.043, 95% CI: [0.02, 0.07]).

**Table 2.**
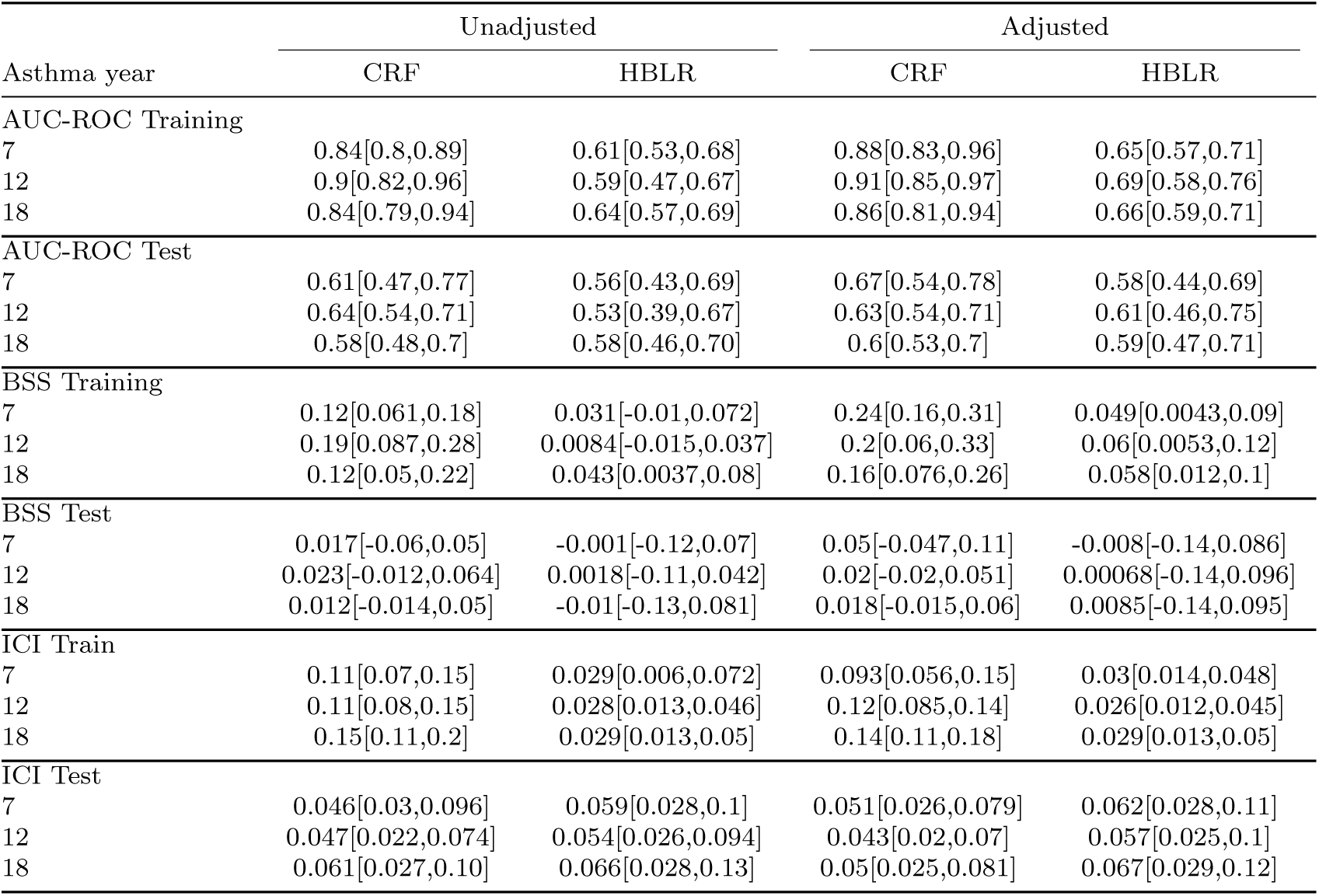
Shows the models performances for Calibrated random forest(CRF) and HBLR across time-points and models. performance is shown with mean[5% CI, 95% CI] and is taken over the 25 test/train splits.

### Predictive Features and Interaction Analysis

To isolate the specific drivers of childhood and adolescent asthma and evaluate potential synergistic effects, the results of the HBLR and the CRF models were evaluated concurrently (Figures 1 and 2).

**Fig. 1.**
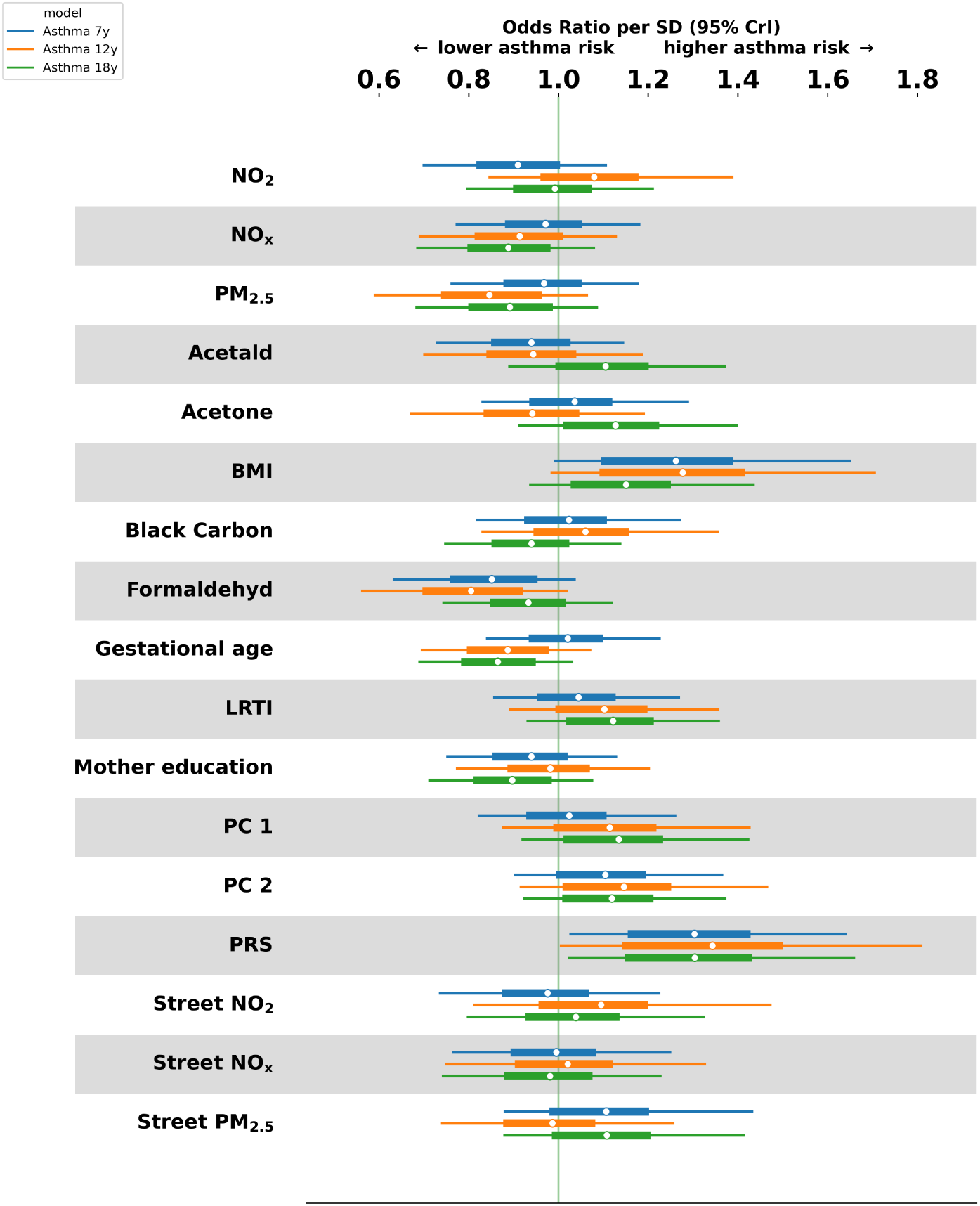
Variable importance from the HBLR at the 7-, 12- and 18-year endpoints, adjusted models. Forest plot of odds ratios per standard deviation increase for asthma at 7 years (blue), 12 years (orange) and 18 years (green). Points denote posterior medians and horizontal bars the 95% HDI; the vertical green line marks the null effect (OR= 1).

**Fig. 2.**
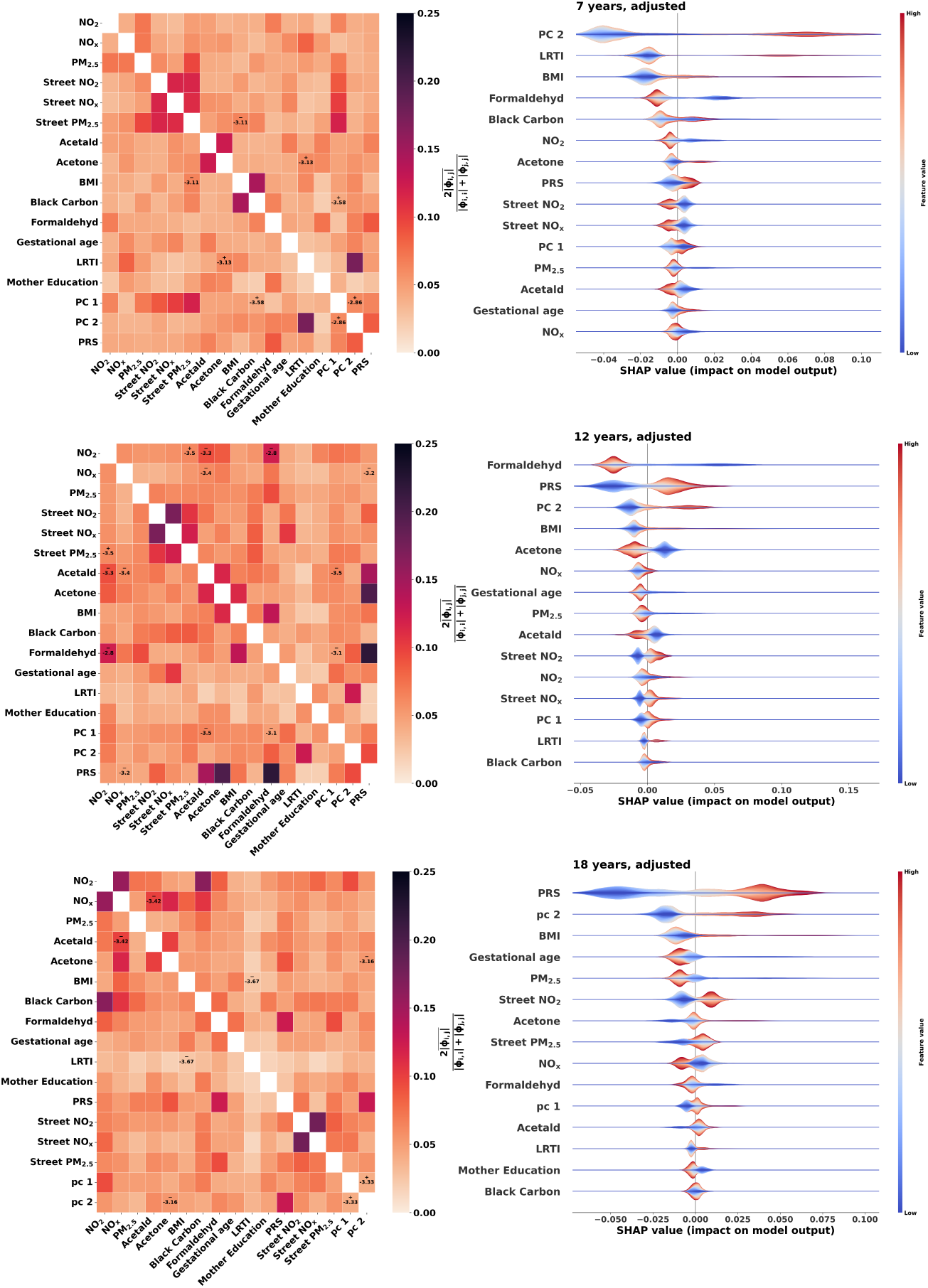
SHAP interpretation of the covariate-adjusted asthma models at 7 years (top row), 12 years (middle row), and 18 years (bottom row). Left: Heatmaps of the pairwise Shapley interaction values; the color bar is scaled from 0.00 to 0.25 to reflect absolute normalized interaction strength. The log_10_ of the raw effect is reported directly within a cell if sign consistency over the validation folds exceeded 80% and the correlation is below 0.3. **Right:** Layered violin plots of out-of-fold SHAP values for the top fifteen features with the largest mean absolute SHAP value, ordered from highest to lowest overall importance. Color encodes feature value intensity (red: high, blue: low); separation across the vertical zero line indicates the direction in which a feature influences the predicted probability of an asthma diagnosis. ^19^

Across both modeling frameworks, genetic susceptibility and physical development emerged as the most dominant and stable predictors of asthma risk up to 18 years. In the HBLR model (Figure 1), the polygenic risk score (PRS) was the only feature whose 95% Highest Density Interval (95% HDI) excluded the null threshold (OR = 1.0) across all three longitudinal endpoints, demonstrating a robust, positive association with asthma at age 7, 12, and 18 years. Conversely, primary particulate and gaseous indoor and outdoor pollutants-including fine particulate matter (PM_2.5_), black carbon, nitrogen oxides (NO*_x_*, NO_2_), and street-level ambient exposures-consistently exhibited posterior median odds ratios close to unity, with all corresponding 95% HDI including OR = 1.0.

The out-of-fold Shap values from the CRF confirmed these structural trends while capturing non-linear relationships (Figure 2, right panels). For the 7-year and 12-year endpoints, feature importance rankings were consistently led by a subset consisting of the PRS, BMI, PC2, and indoor formaldehyde. In alignment with the Bayesian odds ratios, high values of indoor formaldehyde (represented in red) carried negative SHAP values, shifting the model outputs toward decreased predicted probabilities of asthma risk.

The pairwise Shapley interaction heatmaps (Figure 2, left panels) revealed limited high-order interactions among the exposures with consistently weak effect on asthma with an effect size of -4 to -2.8 on log10 scale.

### Sources of indoor air pollution

**Fig. 3.**
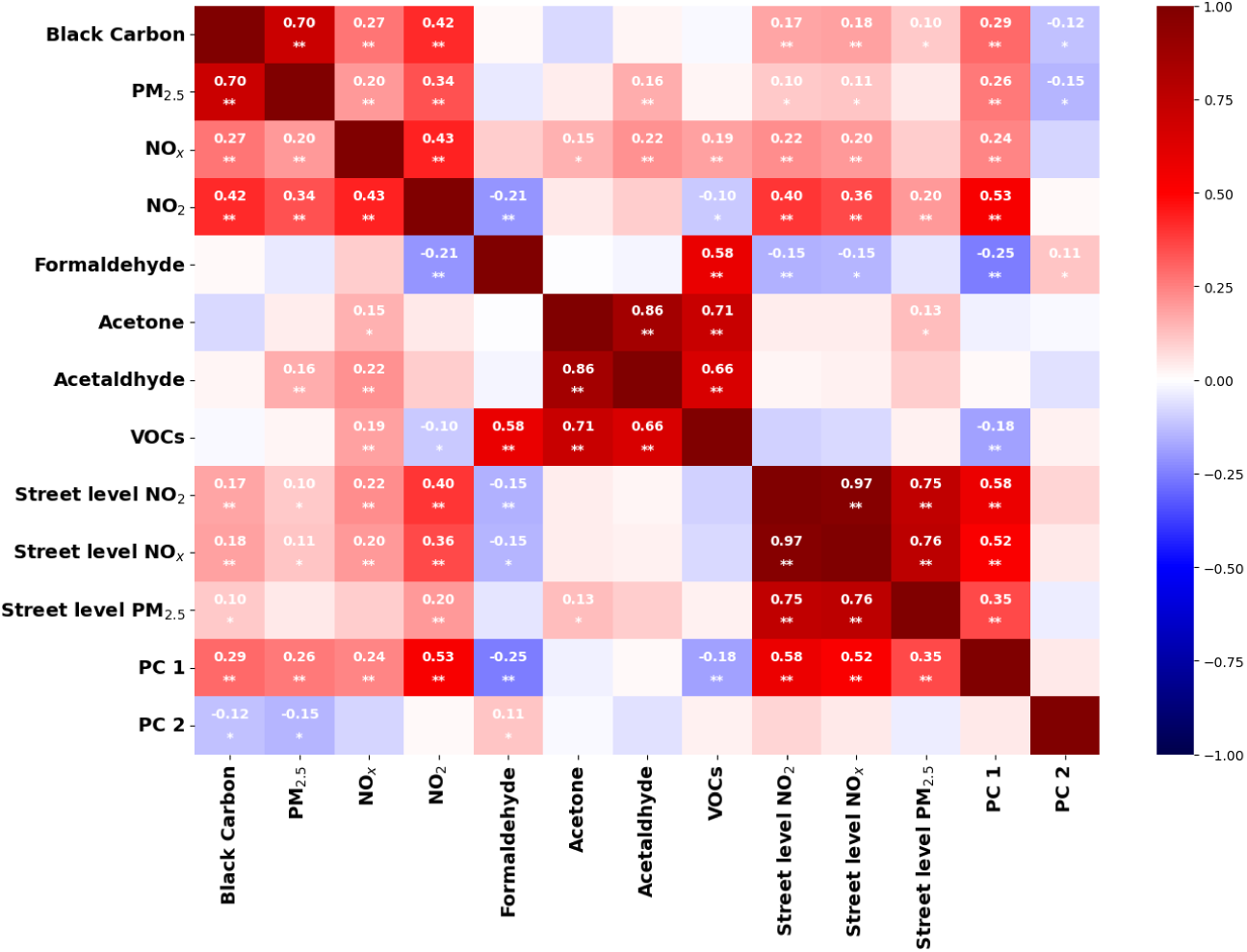
Pairwise Spearman correlation matrix among indoor pollutants (black carbon, PM_2.5_, *NO_x_*, *NO*_2_, formaldehyde, acetone, acetaldehyde, total VOCs), street-level pollutants (NO_2_, NO*_x_*, PM_2.5_) and the two principal components, after KNN imputation (*k* = 25). Annotations mark *^∗^p <* 0.05 (nominal) and *^∗∗^q <* 0.05 (Bonferroni-corrected). Combustion-derived pollutants form a tightly correlated block (*r >* 0.4), and street-level *NO*_2_ and *NO_x_*are nearly collinear (*r* = 0.97)

To characterize the underlying exposure matrix and evaluate spatial patterning, pairwise Spearman correlation coefficients (*r_s_*) were computed across indoor pollutants, street-level modelled values, PC1 and PC2. Combustion-derived indoor particles and gases formed a tightly coupled exposure cluster; specifically, indoor black carbon exhibited robust positive associations with indoor PM_2.5_ (*r_s_* = 0.70, *q <* 0.05) and indoor NO_2_ (*r_s_* = 0.42, *q <* 0.05). At the ambient level, street-level NO_2_ and street-level NO*_x_* demonstrated near-collinearity (*r_s_* = 0.97, *q <* 0.05), and both were strongly bound to the first land-use component (PC 1: *r_s_* = 0.58 and *r_s_* = 0.52, respectively, *q <* 0.05), which primarily tracks traffic-dense and built-up areas. Furthermore, ambient traffic signals clearly penetrated the home, evidenced by the significant correlation between street-level NO_2_ and indoor NO_2_ (*r_s_* = 0.40, *q <* 0.05). In stark contrast, indoor volatile organic compounds (VOCs) exhibited an independent exposure profile. Indoor formaldehyde demonstrated a distinct inverse relationship with traditional traffic markers, correlating negatively with indoor NO_2_ (*r_s_* = −0.21, *q <* 0.05) and the urban gradient component PC 1 (*r_s_* = −0.25, *q <* 0.05). Concurrently, formaldehyde was positively correlated with total VOCs (*r_s_* = 0.58, *q <* 0.05), while acetone and acetaldehyde formed an independent, highly collinear metabolic block (*r_s_* = 0.86, *q <* 0.05).

## Discussion

Baseline descriptive characteristics of the COPSAC_2000_ cohort, stratified by lifetime asthma status, are presented in Table 1. Of the 411 children included in the cohort, 141 (34%) received an asthma diagnosis at least once between ages 0 and 18 years. Body Mass Index (BMI) at age 6 and cooker-hood use were the only covariates that exhibited nominal statistically significant differences between the asthmatic and non-asthmatic groups (*p <* 0.05). Variables displaying marginal differences (*p <* 0.10) included residential floor area and the second principal component (PC2). While tree-based models and HBLR were implemented to evaluate predictive performance, the Shapley Additive Explanations (SHAP) interaction analyses depicted in Figure 2 revealed minimal interactive effects. Due to small estimated effect sizes and limited sample constraints, specific higher-order interactions may have remained below the threshold of detection. Notably, across the 7-year and 12-year longitudinal endpoints, the SHAP feature importance plots identified a consistent top-tier subset of predictive features, specifically the polygenic risk score (PRS), indoor formaldehyde, PC2, and acetone.

### Artifacts and patterns

The absence of a credible pollutant effect in this cohort contrasts with several large-scale studies that have reported associations between ambient PM_2.5_ or NO_2_ and incident childhood asthma. However, most positive findings come from cohorts in which ambient concentrations were substantially higher than those observed in COPSAC_2000_ (median street-level PM_2.5_ of 11.9 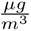 and NO_2_of 21.5 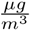). For example, North American or East Asian birth cohorts where associations have been detected. This is consistent with a dose-response relationship in which the exposure contrast in a low-pollution setting is simply too narrow to generate a detectable signal against the background of genetic and contextual variation. The dominance of the polygenic risk score at all three endpoints aligns with recent work showing that genetic architecture accounts for a substantial share of asthma heritability independently of measured environmental exposures [14]. The correlation structure further contextualises the null pollutant finding: combustion-derived indoor species clustered tightly (black carbon and PM_2.5_ had a correlation of *r* = 0.70 and street NO_2_-NO*_x_* of *r* = 0.97, indicating that any single pollutant serves as a proxy for the broader combustion mixture rather than acting independently. Conversely, indoor VOCs especially formaldehyde were statistically independent of the combustion block and inversely correlated with traffic markers (formaldehyde-indoor to NO_2_ had a correlation of *r* = −0.21), reflecting distinct emission sources (building materials, furnishings) that operate on different spatial and temporal scales.

### Simplicity vs non-linearity

The two modelling frameworks were chosen to bracket the plausible complexity of the exposure-outcome relationship. The HBLR assumes a log-linear, additive structure and provides directly interpretable odds ratios with uncertainty quantification through HDI. The calibrated random forest relaxes the linearity and additivity assumptions and, CRF permits detection of higher-order interactions without pre-specification. That both approaches returned near-null pollutant effects posterior ORs spanning unity in the Bayesian model, and negligible mean SHAP values for pollutants in random forest, strengthens the conclusion that the null finding is not an artefact of model misspecification. The modest advantage of the random forest in AUC-ROC (0.58-0.67 versus 0.53-0.61 for HBLR) was not accompanied by meaningful improvement in Brier Skill Score, suggesting that the additional flexibility captured residual non-linearity in covariates (principally BMI and PC2) rather than in the pollutant terms themselves. Overfitting in the tree-based models (training AUC-ROC up to 0.90) further confirms that the feature space does not contain a reproducible pollutant signal strong enough to generalise across folds. The near-zero BSS values (range −0.010 to 0.050) indicate that neither model improved meaningfully over a prevalence-only baseline, reinforcing the interpretation that early-life pollutant exposures at the levels observed do not carry sufficient prognostic information to shift individual risk predictions. Low ICI values (0.043-0.067) confirm that the predicted probabilities were well calibrated in absolute terms; but simply uninformative beyond baseline prevalence.

### Predicted Interaction

We hypothesised that gene-environment and exposure-exposure interactions would amplify the risk conferred by individual pollutants. The pairwise Shapley interaction heatmaps provided little support for this hypothesis: interaction effect sizes ranged from 10^−4^ to 10^−2.8^ on the absolute scale, and no gene-pollutant pair reached a level that would materially alter the predicted probability of asthma. While sign consistency exceeded 80% for a small number of PRS-pollutant pairs, the magnitude of these interactions was negligible relative to the main effects of the PRS itself. This does not rule out the biological plausibility of gene-environment interactions in asthma; pollutant-driven oxidative stress and epigenetic modulation of airway inflammation are well-documented mechanisms [16, 17]. However, it does indicate that such interactions, if present, are either too small to detect in a cohort of this size or require exposure levels above those observed in this Danish setting.

Significant gene-environment interactions have been reported in other childhood cohorts. In the Canadian CHILD study, Choi et al. [18] found that a four-locus genetic risk score interacted with first-year NO_2_ exposure (*p <* 0.01) to modulate risk of recurrent wheeze and asthma, in a sample of approximately 2,500 children. Ambient NO_2_ concentrations in that cohort (estimated by city-specific land-use regression across Vancouver, Edmonton, Winnipeg and Toronto) overlap substantially with those in COPSAC_2000_, suggesting that the discrepancy is driven less by exposure range than by statistical power: the CHILD cohort had roughly six times our sample size. By contrast, Zhu et al. [20] analysed approximately 300,000 UK Biobank participants and found that PM_2.5_, NO_2_ and NO*_x_* each increased adult-onset asthma risk, but identified an additive interaction with lifestyle factors only, not with genetic risk. Notably, the UK Biobank ambient concentrations (mean PM_2.5_ : 9.9 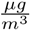; mean NO_2_ : 26.2 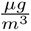) are comparable to those in our cohort, and the null gene-pollution interaction aligns with our own finding. Similarly, Po et al. [19] reported only borderline evidence for a PRS × NO_2_ interaction (coefficient 0.23, 95 % CI: −0.03 to 0.49) in 1,794 children from the Southern California CHS, despite substantially higher ambient concentrations and roughly four times our sample size. That a larger, higher-exposure paediatric cohort yielded an interaction estimate indistinguishable from zero reinforces our conclusion that gene-pollutant synergies, if present at the exposure levels observed here, fall below the detection threshold of cohorts of this scale.

The discrepancy between the Canadian finding and the null or borderline results in the UK Biobank, the Southern California CHS and our own cohort likely reflects differences in the genetic instruments used (4 loci in Choi et al. versus a literature-based PRS in Po et al. versus 212 loci in Zhu et al. versus 23 childhood-onset loci in our study), in the age of onset studied (childhood versus adult), and in the definitions of interaction tested (multiplicative versus additive). Narrower exposure contrasts further reduce statistical power to detect multiplicative effects, and PRS constructed from different variant sets may capture different components of genetic liability. Our results are consistent with the null gene-environment findings from settings with comparable ambient concentrations, extending them with a dual-modelling framework that explicitly tests for non-linear interactions, albeit in a smaller and genetically less diverse sample.

The tight correlation structure among combustion-derived pollutants also limits the ability to attribute interaction effects to specific pollutant pairs, as collinear features can produce spurious Shapley interaction values even under a purely additive model, a concern we addressed by filtering on |*ρ_ij_*| ≤ 0.3 and requiring sign consistency *C_ij_*≥ 0.8 on validation folds.

### Strengths and limitations

Indoor PM_2.5_ and black carbon were measured over only 10 days per visit, and the VOCs over approximately 10 weeks, so even after flow correction and periodic Gaussian-process seasonal adjustment, residual measurement error remains a source of uncertainty [13]. Bedroom-based sampling does not capture daycare, school or outdoor micro-environments. BMI at 6 years was missing for 109 of 411 children (26%) and was imputed within folds; The cohort was recruited from mothers with physician-diagnosed asthma, enriching for genetic risk and limiting direct generalisation to general populations. Finally, with 411 children and at most 141 asthma cases, we cannot exclude weak gene-environment interactions or distinguish individual pollutant associations within tightly correlated blocks. We therefore constrained model complexity (hierarchical shrinkage and a small curated random forest, with no mixture-method layer) to match the information available rather than estimate effects the cohort cannot resolve. Strengths include a well-defined, physician-confirmed asthma diagnosis at 7, 12 and 18 years, prospectively assessed in a single-centre setting. Indoor PM_2.5_ and black carbon were measured by trained staff, and the passive samplers required only shipping and start/stop handling by parents. We applied a dual modelling approach, using both a CRF and a HBLR, to obtain interpretable odds ratios, calibrated risk estimates, and some purchase on the correlation structure. Measurements were corrected for both pump-flow drift and seasonal sampling bias, and both corrections were selected by household-level train/test evaluation and are outcome-blind.

## Conclusion

In this prospectively followed Danish birth cohort with repeated indoor measurements corrected for pump-flow drift and seasonal sampling bias, neither cumulative early-life pollutant exposure nor gene–environment interactions were associated with asthma at 7, 12 or 18 years of age. Both a HBLR and a calibrated random forest converged on near-null pollutant effects, with the polygenic risk score as the only predictor whose credible interval excluded the null at all three endpoints. The improved exposure correction did not unmask a previously obscured association, suggesting that the null finding reflects a genuinely narrow exposure contrast rather than measurement artefact. Larger, more diverse cohorts with higher ambient concentrations are needed to resolve whether pollutant-specific or interactive effects emerge when the exposure range is wider.

## Supporting information

Supplemental sections

## Data Availability Statement

The clinical and omics data utilized in this study are derived from the prospective COPSAC clinical birth cohorts. Due to the sensitive nature of the human participant data and strict ethical and legal constraints, the datasets are not publicly archived in an open repository. De-identified data may be made available to qualified researchers upon reasonable request to the corresponding author, subject to approval by the COPSAC steering committee and the relevant Danish data protection authorities.

## Code Availability

Code and synthetic data are available in the GitHub repository at https://github.com/MichaelForsmann/Predicting-asthma-from-Early-life-Cumulative-indoor-and-outdoor-exposures

## Acknowledgements

We express our deepest gratitude to the children and families of the COPSAC_2000_ cohort study for their support and commitment. We acknowledge and appreciate the unique efforts of the COPSAC research team. We would like to acknowledge the work of the late Professor Hans Bisgaard, founder of COPSAC and head of the clinical research centre for more than 25 years. Hans was a dedicated, innovative physician-scientist who pushed the field of asthma research forward. He contributed immensely to paediatric research through the COPSAC birth cohorts, as well as through a large number of other clinical studies on childhood asthma. His work and ideas live on in the studies conducted in the birth cohort. Thank you for being a great inspiration to us all.

## Author Contributions

Conceptualization, methodology, visualization, formal analysis and investigation: Michael Forsmann. Original draft preparation and writing, review and editing: Michael Forsmann, Mario Lovrić and Morten Arendt Rasmussen. Data collection, review and editing: Michael Forsmann, Frederikke R. Skov. Review and editing: Frederikke R. Skov, Olalekan Akintande, Jakob Stokholm, Klaus Bønnelykke and Ann-Marie Malby Schoos. Funding acquisition: Mario Lovrić. Supervision: Morten Arendt Rasmussen. All authors read and approved the final manuscript.

## Ethics Approval and Consent to Participate

The study protocol was approved by the Ethics Committee of Copenhagen (KF 01-289/96) and the Danish Data Protection Agency (2002-41-2434), and written consent was obtained from the parents of the participating infants.

