## Supplemental sections for "Associations Between Early-Life Indoor and Outdoor Air Pollution Exposure and Childhood Asthma"

This file contains supplementary methods, figures and tables supporting the main manuscript: exposure-correction procedures for pump-flow drift and seasonal sampling bias, imputation diagnostics, the modelling pipeline, model-performance distributions, random-forest SHAP attributions, the pollutant exposure distribution and asthma Diagnosis.

#### S1 Adjustment for pump flow for black carbon and PM$_{2.5}$

$PM_{2.5}$ and black carbon were measured in the children's bedroom over a period of 10 days at 6, 12, and 18 months of age. Trained field staff initiated and terminated each measurement session, recording the pump flow rate at both the start and the end of the sampling period. Measurements were performed using a KTL $PM_{2.5}$ cyclone coupled with a BGI400 pump [1, 2].

Reflectance of the collected filters provides a measure of their blackness, which serves as an indicator of the black carbon content of the sample. The sampling campaigns were designed to cover a 10-day period per household; however, practical constraints introduced variation in both the actual measurement duration and the age of the children at the time of sampling. In addition, seasonal variation in ambient pollution levels adds a further source of heterogeneity across households, which is addressed separately in Supplementary Section S2 .

The measured concentration depends on the pump flow rate, since the effective aerodynamic cut-point of the cyclone ($D_{cut50}$) decreases as flow increases. A lower cut-point excludes a larger fraction of the coarse tail of the size distribution, so the collected mass, and hence the reported concentration, falls systematically. The relationship between the cut-point and the operating parameters is given by

$$D_{cut50} = \sqrt{\frac{9\,\mu\,W}{2\,\pi\,N_e\,v_i\,(\rho_p - \rho_g)}} \tag{S1}$$

Here $D_{\mathrm{cut50}}$ is the 50% separation cut-off diameter of the cyclone separator in metres, $\mu$ the dynamic viscosity of the carrier gas in $\mathrm{kg\,m^{-1}\,s^{-1}}$, and $W$ the characteristic width of the cyclone inlet in metres. The particle and carrier-gas densities are denoted $\rho_p$ and $\rho_g$ respectively, both in $\mathrm{kg\,m^{-3}}$, while $N_e$ is the dimensionless effective number of turns within the cyclone body and $v_i$ the inlet gas velocity in $\mathrm{m\,s^{-1}}$. Of these, only $v_i$ varies across measurements in the present setting, and it does so in direct proportion to the pump flow, which is what makes the flow rate the operative source of measurement bias.

Because the ambient particle size distribution varies between households and over time, reflecting different emission sources, ventilation behaviour and meteorological conditions, the mapping from pump flow to measurement bias is household-specific and cannot be characterised analytically in a clinical cohort setting. We therefore adopted a data-driven approach using Gaussian process (GP) regression. Flow was recorded at both the start and the end of each session, and the end-of-sampling value was used for the correction, since pump drift accumulates over the 10-day deployment and the final reading therefore best characterises the cut-point in effect over the majority of the sampled period.

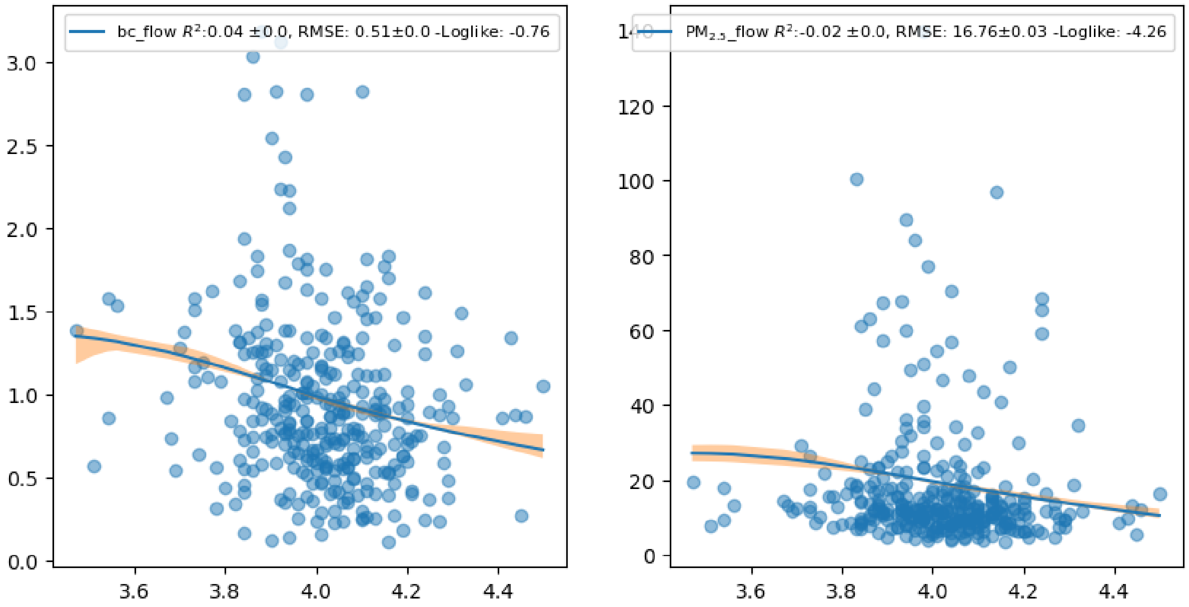

Figure S1 : Empirical dependence of measured indoor concentrations on the end-of-sampling pump flow rate ($\mathrm{l\,min^{-1}}$). **Left:** black carbon; **right:** $\mathrm{PM}_{2.5}$. Blue points are the raw 10-day household measurements. Both pollutants show a monotonic decrease in measured concentration with increasing flow, consistent with the cut-point shift described in Supplementary Eq. (S1 ). The proportion of variance explained is small ($R^2 = 0.04$ and $-0.02$ respectively), as between-household differences in true concentration dominate the total variation; the correction is motivated by the mechanistic relationship in Supplementary Eq. (S1 ) rather than by the strength of the empirical fit.

As shown in Supplementary Fig. S1 , we fitted a GP with a radial basis function (RBF) kernel to model the dependence of $PM_{2.5}$ and black carbon concentrations on the recorded pump flow. The corrected concentration for observation $i$ is the ratio of the GP prediction at the nominal target flow of $4\,\mathrm{l\,min^{-1}}$ to the prediction at the actual end-of-sampling flow $f_i$:

$$C_{i,\mathrm{adj}} = \frac{\mathrm{GP}_\theta(l_f,\,\sigma_f,\,4)}{\mathrm{GP}_\theta(l_f,\,\sigma_f,\,f_i)}\,C_i \tag{S2}$$

In this expression $C_i$ is the uncorrected concentration and $C_{i,\mathrm{adj}}$ the flow-adjusted concentration, while $\mathrm{GP}_\theta$ denotes the Gaussian process predictive mean parameterised by length-scale $l_f$ and signal variance $\sigma_f$, evaluated at the nominal flow in the numerator and at the observed flow $f_i$ in the denominator. Being multiplicative and monotonic in $f_i$, the correction preserves the relative ranking of concentrations recorded at a given flow while removing the systematic bias introduced by flow deviations.

As for the seasonal correction, the flow-correction function was selected by train/test evaluation with splitting at the household level, so that no household contributed measurements to both sides. The selected function was then fitted on the full cohort and applied to all measurements. Since it regresses concentration on pump flow and never uses asthma status, the transformation is outcome-blind and cannot transmit outcome information into the held-out folds of the prediction pipeline.

#### S2 Seasonal scaling of samples

This section documents the seasonal correction described in Eq. (1) of the main text. The correction function was selected by train/test evaluation with splitting at the household level, so that no household contributed measurements to both sides. The correction used a Gaussian process with a periodic kernel. The periodic GP was retained for the main analysis because it recovers the seasonal cycle without the boundary artefacts of the four-season step correction (Supplementary Fig. S3 ). The same function family was applied uniformly to all pollutants to keep the corrected values comparable across compounds; for pollutants with little seasonal structure the fitted function is close to constant and the correction reduces to approximately unity, so applying it uniformly costs nothing. The selected function was then fitted on the full cohort and applied to all measurements. Since it regresses concentration on calendar week and never uses asthma status, this transformation is outcome-blind and cannot transmit outcome information into the held-out folds of the prediction pipeline.

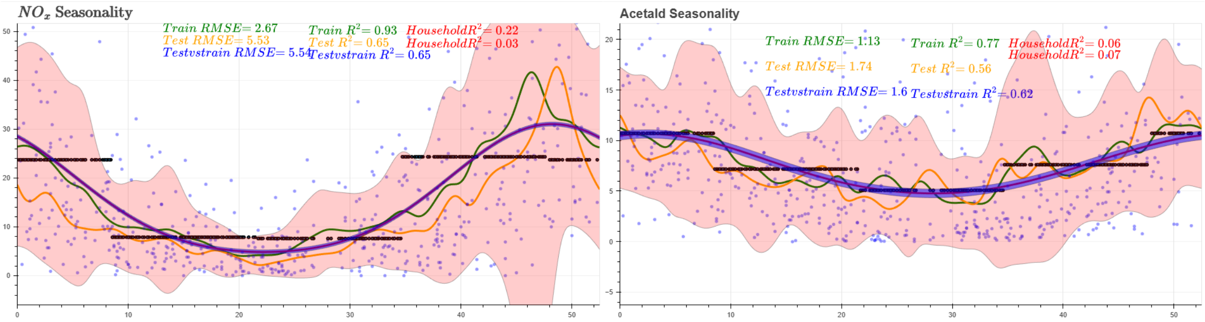

Figure S2 : Periodic Gaussian process seasonal correction function fitted to two indoor pollutants. The periodic correction is shown in blue and the standard (non-periodic) correction in black; the pink envelope denotes the standard deviation of the data and orange points the rolling mean. **Left:** $NO_x$, a highly seasonal combustion-derived pollutant, for which the periodic kernel improves the explained household variance from $R^2_{\text{old}} = 0.03$ to $R^2_{\text{new}} = 0.22$. **Right:** acetaldehyde, dominated by indoor cleaning and ventilation sources and exhibiting only weak seasonal dependence; here the periodic and simple corrections converge and the seasonal function reduces effectively to a constant, so the correction leaves the measurement essentially unchanged.

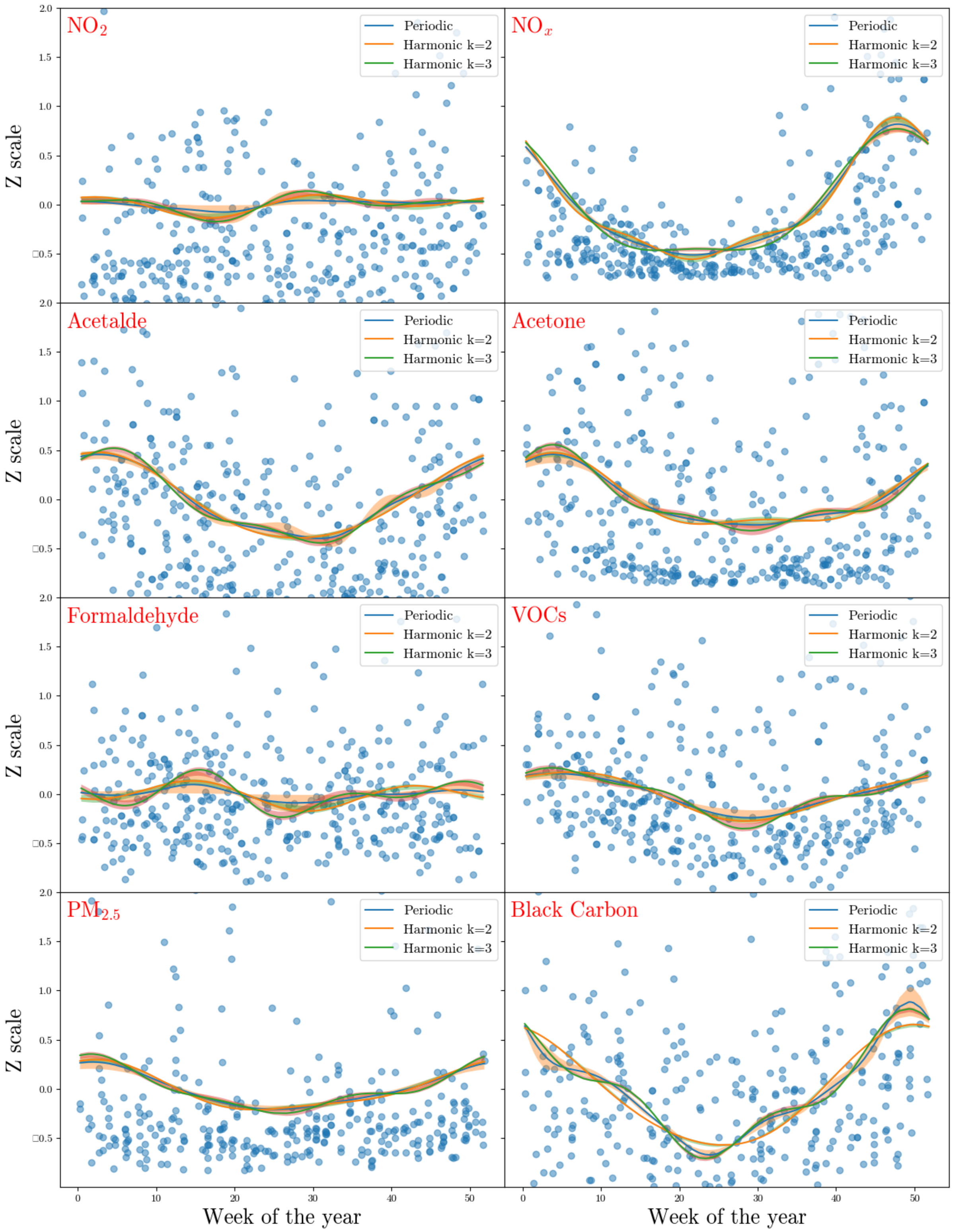

Figure S3 : Per-pollutant comparison of the seasonal-function families considered for the correction in Eq. (1) of the main text. For each of the eight pollutant series ($NO_x$, $NO_2$, acetaldehyde, acetone, formaldehyde, total VOCs, $PM_{2.5}$ and black carbon) the panels show the raw household-level measurements (blue points), the unadjusted yearly rolling mean (dark line), harmonic regression fits with 2 and 3 terms, and the periodic Gaussian-process fit used in the main analysis. Envelopes in the same colour show one standard deviation of the periodic GP uncertainty.

### S3 Pollutants versus sink and sources

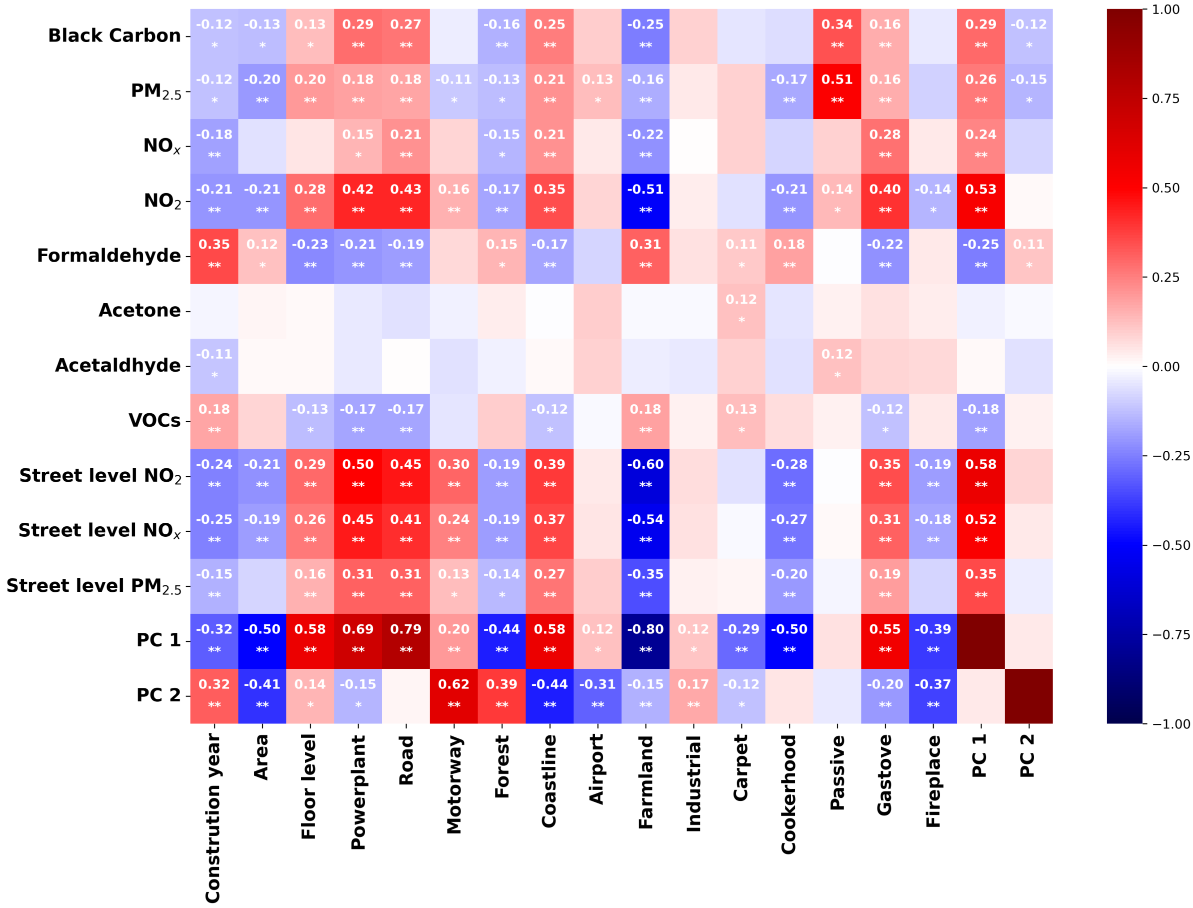

Figure S4 : Spearman correlation matrix between indoor and street-level pollutants (rows) and indoor sinks/sources, building characteristics, and the two PCs (columns). Only statistically significant cells are annotated: $^*p < 0.05$ (nominal) and $^{**}q < 0.05$ (Bonferroni corrected). Colour intensity reflects the magnitude of the correlation, with dark red/orange indicating strong positive and dark blue strong negative associations.

### S4 Imputation via $k$-nearest neighbours and correlation structure

Missing values in the exposure and covariate data were imputed using a $k$-nearest neighbour (KNN) approach, in which each incomplete observation was filled from the $k$ nearest observations in the training fold under a uniform weighting. The number of neighbours was set to $k = 25$ after evaluating imputation quality across $k \in \{5, 10, 15, 20, 25\}$. Figure 3 in the main text displays the pairwise Spearman correlations among the indoor and outdoor air pollution variables, selected lifestyle factors (e.g., parental education level), and health-related covariates (polygenic asthma risk score, history of lower respiratory tract infections, and gestational age). The correlation matrix confirms the expected positive associations between combustion-related pollutants (e.g., $PM_{2.5}$, black carbon, $NO_2$); it also illustrates why KNN is a strong candidate for imputation, since many of the variable pairs are significantly correlated after Bonferroni correction.

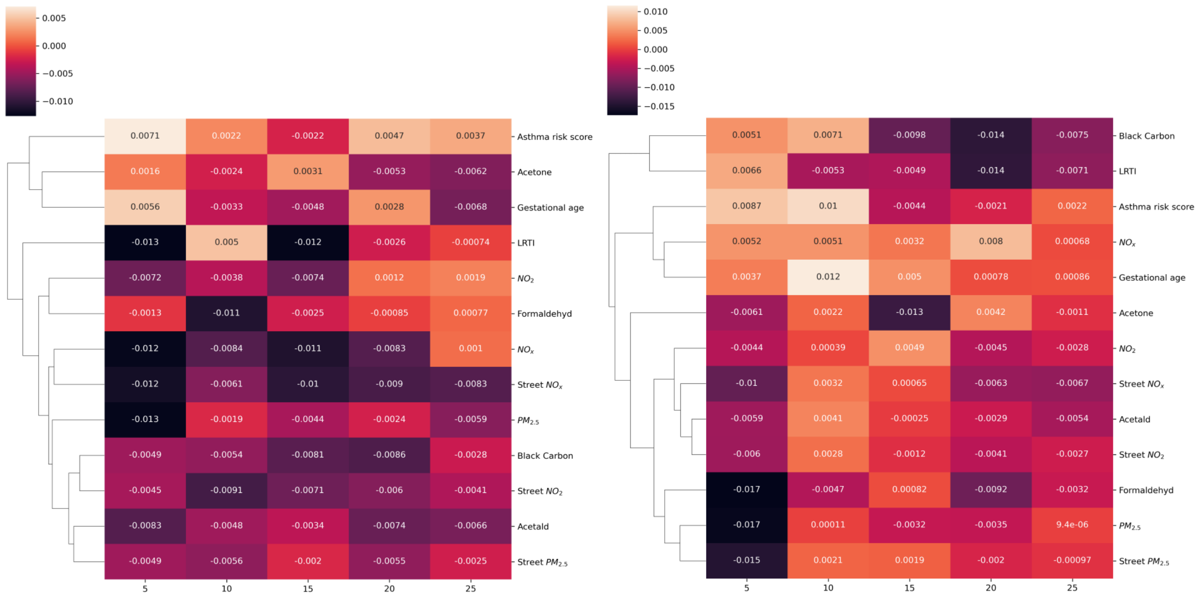

Figure S5 : Sensitivity of the KNN imputation to the number of neighbours $k \in \{5, 10, 15, 20, 25\}$, adjusted feature set. **Left:** mean absolute change in pairwise Spearman correlation between exposures, sinks, sources and clinical covariates relative to the complete-case correlation matrix. **Right:** mean absolute change in marginal feature values (imputed minus complete-case median, on the Z-scaled scale). Both panels show small, near-zero deviations across all $k$; $k = 25$ was retained for the main analysis as the value minimising the joint deviation.

To assess the robustness of the imputation, we compared the marginal distributions and pairwise correlation structures before and after imputation. Supplementary Fig. S6 and Supplementary Fig. S5 present these comparisons for the unadjusted and adjusted feature sets respectively. In both, the mean absolute change in pairwise Spearman correlation and in marginal feature values relative to the complete-case data is near zero across all values of $k$, indicating that the KNN imputation does not introduce spurious associations or materially shift the marginals.

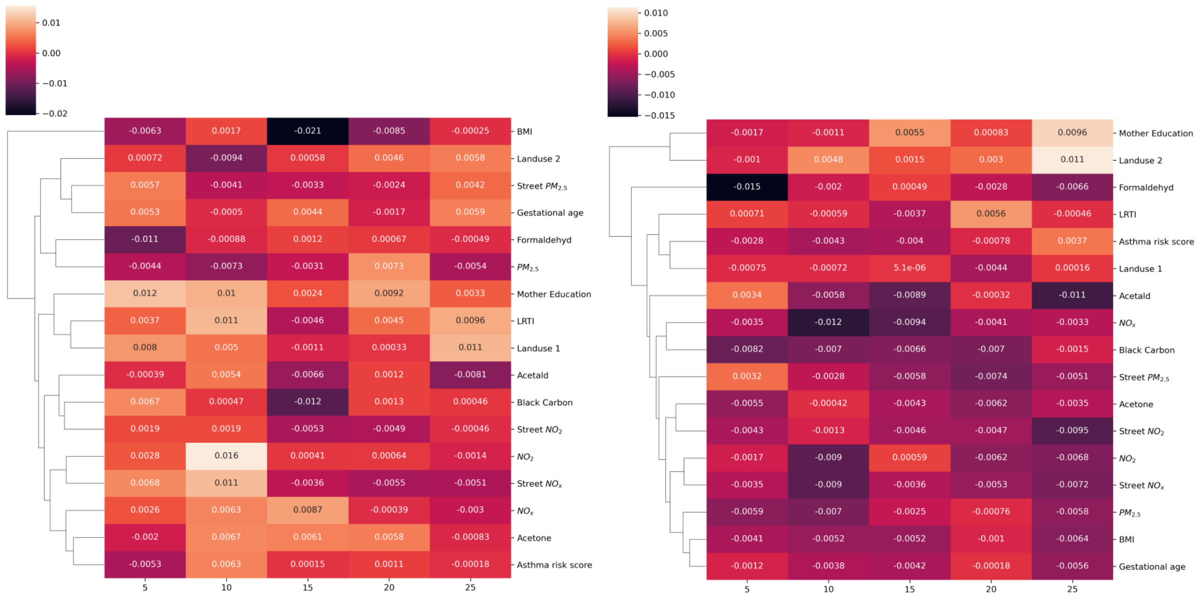

Figure S6 : Sensitivity of the KNN imputation to the number of neighbours $k \in \{5, 10, 15, 20, 25\}$, unadjusted feature set. **Left:** mean absolute change in pairwise Spearman correlation between exposures, sinks, sources and clinical covariates relative to the complete-case correlation matrix. **Right:** mean absolute change in marginal feature values (imputed minus complete-case median, on the Z-scaled scale). Both panels show small, near-zero deviations across all $k$; $k = 25$ was retained for the main analysis as the value minimising the joint deviation.

### S5 Calibrated random forest SHAP values

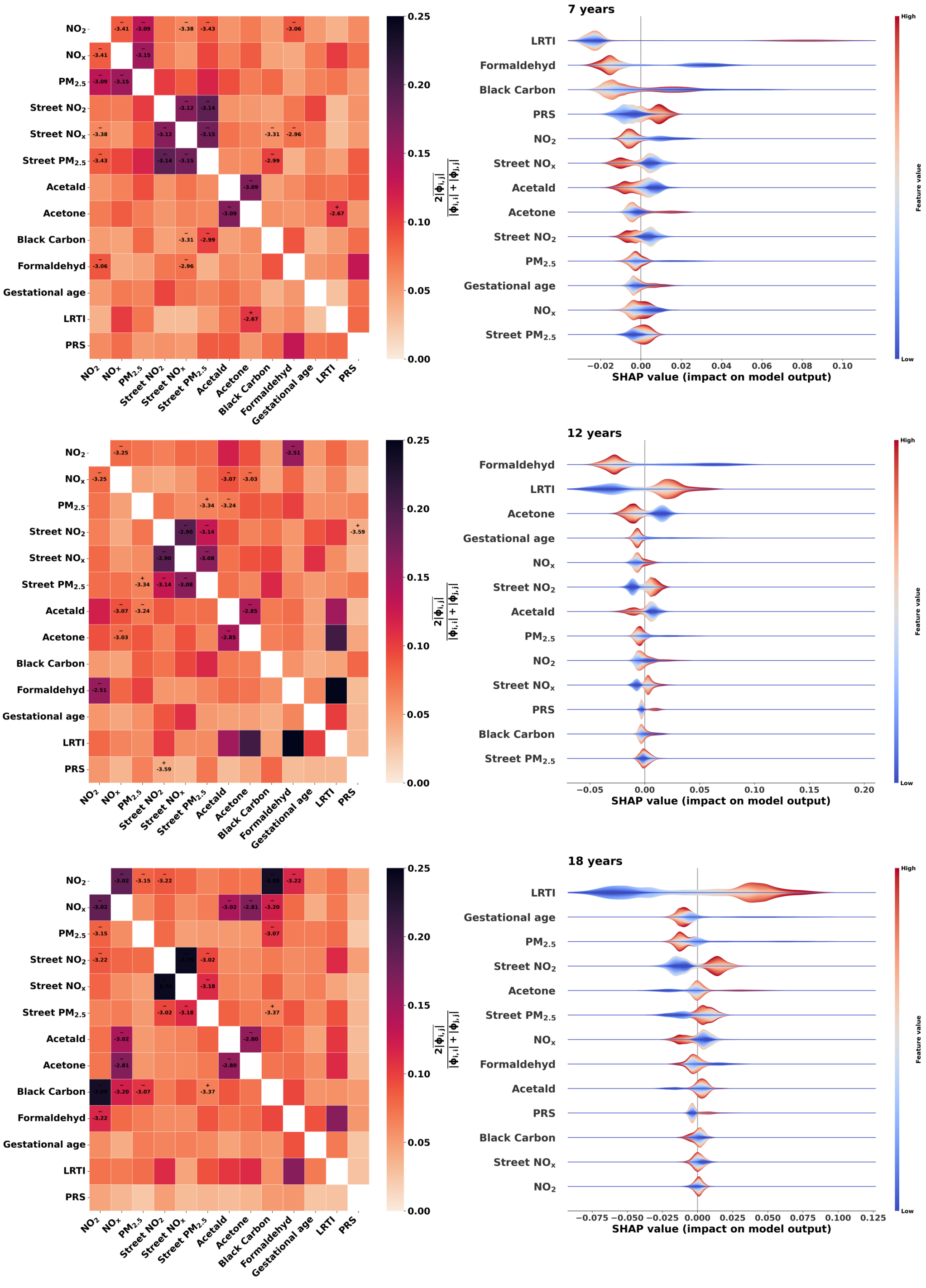

Figure S7 : SHAP value beeswarm plots for the **unadjusted** calibrated random forest predicting physician-confirmed asthma at **7, 12 and 18 years** (top to bottom). Each point represents one child; horizontal position gives the SHAP contribution to the predicted log-odds and colour encodes the standardised feature value (red = high, blue = low). Features are ordered top-to-bottom by mean absolute SHAP value within each panel. At 7 years the cumulative LRTI count ranks first, followed by indoor formaldehyde and black carbon; at 12 years indoor formaldehyde ranks first, followed by the cumulative LRTI count, acetone and the street-level $NO_2$ and $NO_x$ terms; at 18 years the cumulative LRTI count ranks first, followed by street-level $NO_2$ and indoor $PM_{2.5}$. Across all three endpoints, high formaldehyde values carry negative contributions.

#### S6 Unadjusted odds ratios HBLR

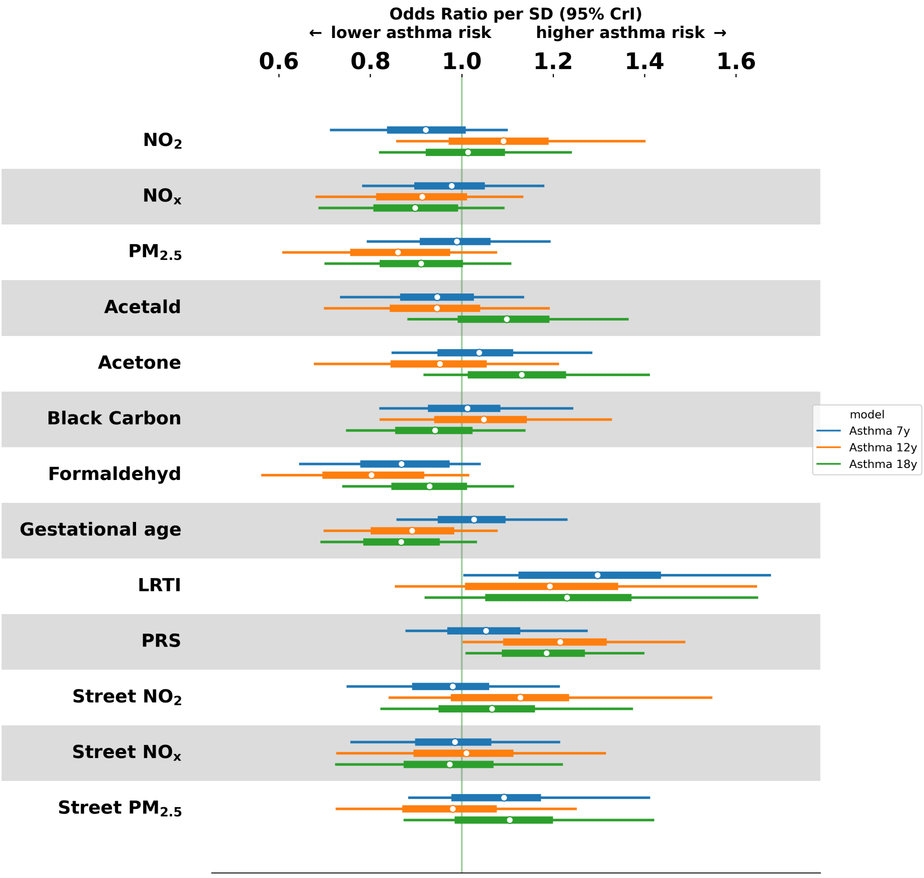

Figure S8 : Variable importance from the **unadjusted** HBLR at the 7-, 12- and 18-year endpoints. Forest plot of posterior odds ratios per standard-deviation increase for asthma at 7 years (blue), 12 years (orange) and 18 years (green). Points denote posterior medians and horizontal bars the 95 % HDI; the vertical line marks the null effect (OR = 1). Unlike the covariate-adjusted analysis in Figure 1 of the main text, these models omit BMI, maternal education and the two land-use principal components.

#### S7 Modelling pipeline

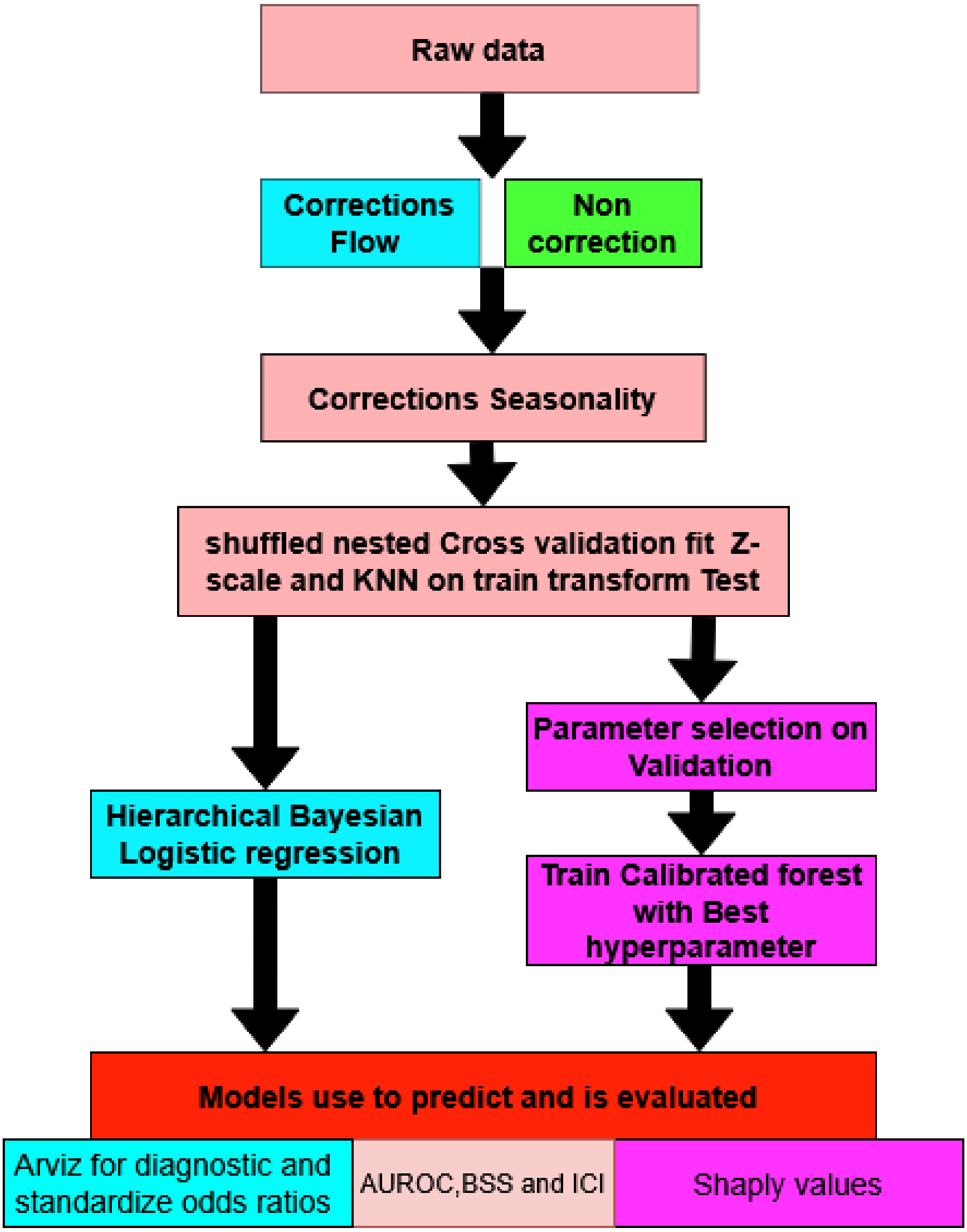

Figure S9 : Modelling pipeline for predicting asthma at ages 7, 12 and 18 years. Raw indoor and outdoor exposures are first corrected for pump-flow drift ($PM_{2.5}$, black carbon) and then for seasonal sampling bias using a periodic Gaussian process. Within each fold of the shuffled stratified nested cross-validation (M= 5 outer, N= 5 inner), Z-scaling and uniform-weighted KNN imputation ($k = 25$) are fitted on the training set and applied to the held-out test set, preventing information leakage. Two complementary models are then trained: a HBLR (left branch), evaluated through ArviZ posterior diagnostics and odds ratios; and a calibrated random forest with Sigmoid regression (right branch), evaluated through SHAP values after hyper-parameter selection on the inner validation folds. Both models are scored on the outer test folds using AUC-ROC, Brier skill score (BSS) and integrated calibration index (ICI).

#### S8 Distribution of performance

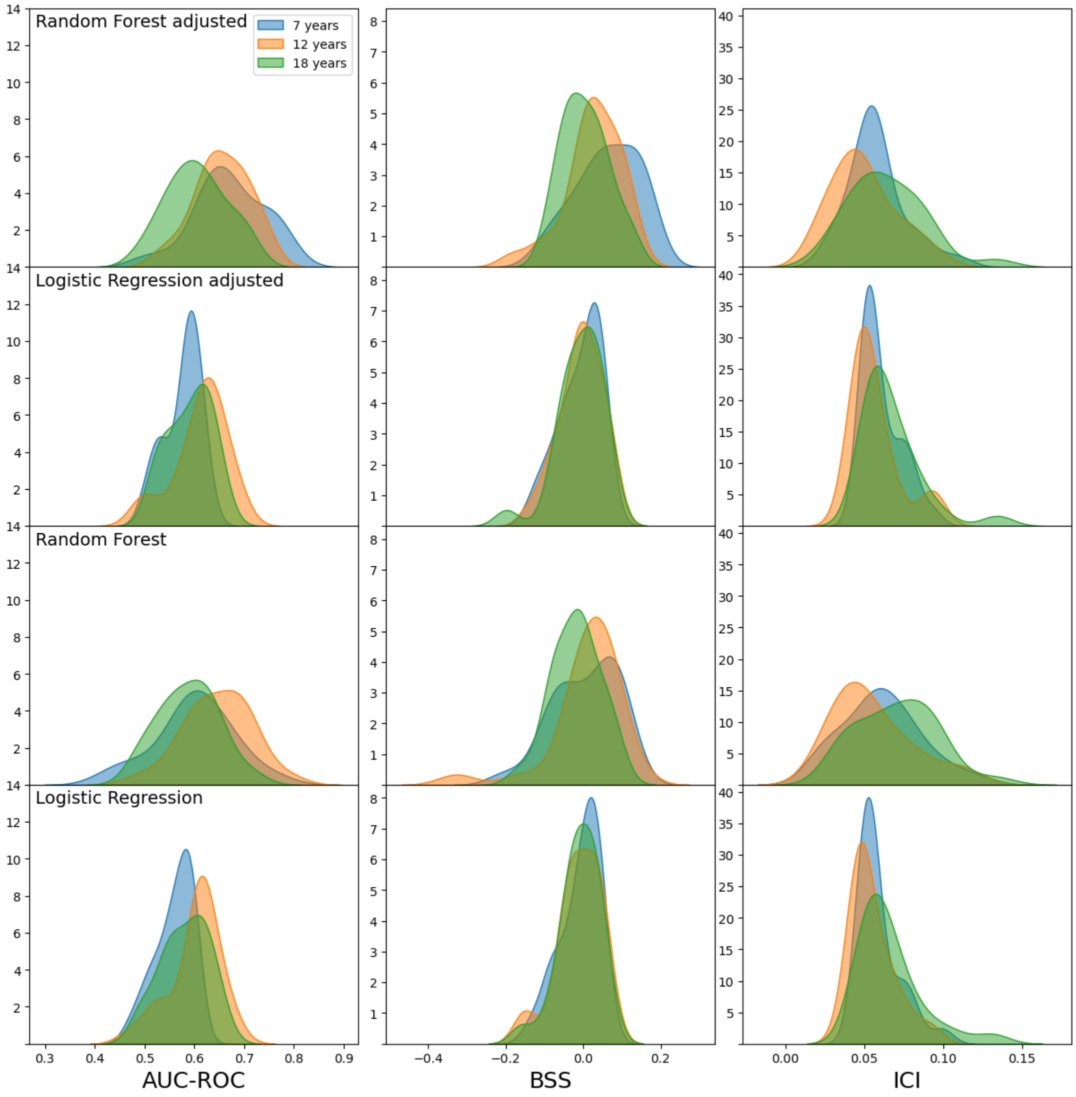

Figure S10 : Distributions of AUC-ROC, BSS and ICI across the outer cross-validation folds for both model classes. **Top two rows:** adjusted models. **Bottom two rows:** unadjusted models. Within each panel the distributions are stacked across the three asthma endpoints (7, 12 and 18 years) and shown separately for the calibrated random forest (RF) and the HBLR (LR)

### S9 Sampled performance for HBLR

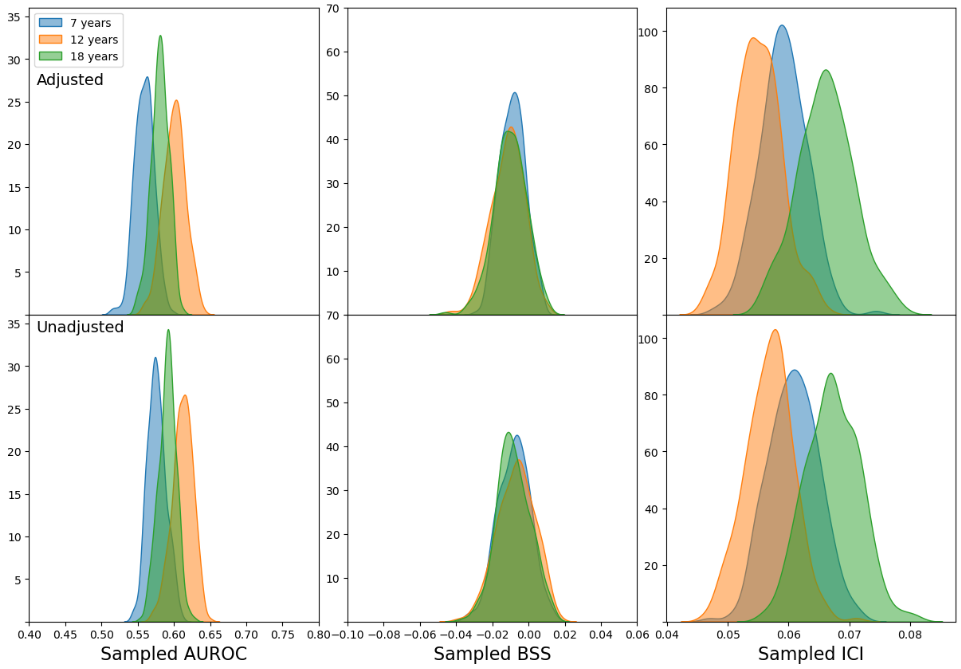

Figure S11 : Posterior predictive distributions of performance metrics for the hierarchical Bayesian logistic regression (HBLR) model across three prediction horizons (7, 12, and 18 years). The top row shows results from covariate-adjusted models and the bottom row from unadjusted models. Three metrics are displayed: discrimination (sampled AUROC; left), overall predictive accuracy relative to a reference model (sampled Brier Skill Score [BSS]; centre), and calibration (sampled Integrated Calibration Index [ICI]; right). Density curves are constructed from posterior samples to characterise uncertainty around each point estimate. AUROC values increase modestly with longer prediction horizons, while BSS values cluster near or below zero, indicating limited improvement over a naïve reference forecast. Lower ICI values reflect better calibration, with the 7-year horizon generally exhibiting the tightest calibration. Differences between adjusted and unadjusted models are small across all metrics and horizons.

### S10 Distribution of pollutants

Table S1 : Distribution of indoor and street-level pollutants. Values shown are the 2.5%, 50% (median) and 97.5% quantiles, and the mean.

| | Indoor concentrations | | | | | | | Street level | | |
|---|---|---|---|---|---|---|---|---|---|---|
| | $NO_x$ | $NO_2$ | Formaldehyde | Acetone | Acetaldehyde | Black carbon | $PM_{2.5}$ | $NO_2$ | $NO_x$ | $PM_{2.5}$ |
| Units | $\mu g\,m^{-3}$ | $\mu g\,m^{-3}$ | $\mu g\,m^{-3}$ | $\mu g\,m^{-3}$ | $\mu g\,m^{-3}$ | $m^{-1}$ | $\mu g\,m^{-3}$ | $\mu g\,m^{-3}$ | $\mu g\,m^{-3}$ | $\mu g\,m^{-3}$ |
| 2.5% | 1.1 | 2.9 | 6.3 | 1.1 | 0.7 | 0.4 | 6.5 | 12.5 | 14.1 | 9.1 |
| 50% | 10.3 | 7.4 | 17.7 | 7.5 | 6.1 | 0.9 | 13.5 | 21.5 | 27.1 | 11.9 |
| 97.5% | 43.4 | 19.9 | 46.3 | 34.6 | 20.5 | 1.8 | 67.5 | 42.2 | 97.5 | 15.3 |
| Mean | 14.0 | 8.4 | 19.4 | 9.6 | 7.1 | 1.0 | 19.2 | 22.2 | 32.8 | 12.0 |

### S11 Asthma diagnosis

The diagnosis was based on: (a) recurrent troublesome lung symptoms, defined as ≥5 episodes each lasting ≥3 consecutive days within 6 months, or daily symptoms for ≥4 consecutive weeks;

(b) typical asthma symptoms (e.g. prolonged nocturnal cough, exercise-induced symptoms, and nighttime awakenings); (c) clinical response to intermittent inhaled short-acting $\beta$-agonist; and (d) response to a 3-month trial of inhaled corticosteroids (ICS) followed by relapse after stopping ICS. Relapse was defined as either $\geq$2 symptom episodes lasting $\geq$3 days within 3 months, or symptoms occurring $\geq$2 times per week for 2 consecutive weeks. Severe acute exacerbations requiring hospitalisation or oral corticosteroids also resulted in an asthma diagnosis and initiation of ICS treatment (typically for 6 months). Asthma remission was defined as absence of asthma symptoms and no need for asthma medication for 12 months. From age 7 years onward, the diagnosis was retrospectively validated according to European Respiratory Society (ERS) criteria [3]. Objective confirmation required fulfilment of $\geq$2 of the following at any clinical visit:

(1) obstructive lung function ($FEV_1 \leq 80\%$ of predicted value, $FEV_1$/FVC-index <80% by spirometry, and/or sRaw >1.6 kPa/s),
(2) positive bronchodilator reversibility ($\geq$12% increase in $FEV_1$ and/or $\geq$30% improvement in sRaw after inhaled $\beta_2$-agonist),
(3) airway inflammation (FeNO $\geq$25 ppb), and/or
(4) bronchial hyperresponsiveness to methacholine ($\geq$20% fall in $FEV_1$ at methacholine dose $\leq$8 mg/mL), exercise ($\geq$10% decrease in $FEV_1$) and/or cold air provocation ($\geq$10% decrease in $FEV_1$) [4, 5].
